# Symptoms compatible with long-COVID in healthcare workers with and without SARS-CoV-2 infection – results of a prospective multicenter cohort

**DOI:** 10.1101/2021.10.19.21265187

**Authors:** Carol Strahm, Marco Seneghini, Sabine Güsewell, Thomas Egger, Onicio Leal, Angela Brucher, Eva Lemmenmeier, Dorette Meier Kleeb, J. Carsten Möller, Philip Rieder, Markus Ruetti, Remus Rutz, Hans-Ruedi Schmid, Reto Stocker, Danielle Vuichard-Gysin, Benedikt Wiggli, Ulrike Besold, Stefan P. Kuster, Allison McGeer, Lorenz Risch, Andrée Friedl, Matthias Schlegel, Dagmar Schmid, Pietro Vernazza, Christian R. Kahlert, Philipp Kohler

**Affiliations:** Cantonal Hospital St Gallen, Division of Infectious Diseases and Hospital Epidemiology, St Gallen, Switzerland; Epitrack, Recife, Brazil; Department of Economics, University of Zurich, Zurich, Switzerland; Psychiatry Services of the Canton of St. Gallen (South), Switzerland; Clienia Littenheid AG, Private Clinic for Psychiatry and Psychotherapy, Littenheid, Switzerland; Kantonsspital Baden, Division of Occupational Health, Baden, Switzerland; Center for Neurological Rehabilitation, Zihlschlacht, Switzerland; Hirslanden Clinic, Zurich, Switzerland; Fuerstenland Toggenburg Hospital Group, Wil, Switzerland; Kantonsspital Baden, Central Laboratory, Baden, Switzerland; Thurgau Hospital Group, Division of Infectious Diseases and Hospital Epidemiology, Muensterlingen, Switzerland; Kantonsspital Baden, Division of Infectious Diseases and Hospital Epidemiology, Baden, Switzerland; Geriatric Clinic St. Gallen, St. Gallen, Switzerland; Federal Office of Public Health, Bern, Switzerland; Sinai Health System, Toronto, Canada; Labormedizinisches Zentrum Dr Risch Ostschweiz AG, Buchs, Switzerland; Private Universität im Fürstentum Liechtenstein, Triesen, Liechtenstein; Center of Laboratory Medicine, University Institute of Clinical Chemistry, University of Bern, Inselspital, Bern, Switzerland; Cantonal Hospital St Gallen, Clinic for Psychosomatic and Consultation Psychiatry, St. Gallen, Switzerland; Children’s Hospital of Eastern Switzerland, Department of Infectious Diseases and Hospital Epidemiology, St. Gallen, Switzerland

**Author notes:** **Corresponding authors:** Carol Strahm, MD, Division of Infectious Diseases and Hospital Epidemiology, Cantonal Hospital St. Gallen, Rorschacherstrasse 95, 9011 St. Gallen, Switzerland. **Alternative corresponding address:** Philipp Kohler, MD MSc, Division of Infectious Diseases and Hospital Epidemiology, Cantonal Hospital St. Gallen, Rorschacherstrasse 95, 9011 St. Gallen, Switzerland.

**Keywords:** Long-COVID, serology, asymptomatic, healthcare workers, risk factors

## Abstract

**Background:** The burden of long-term symptoms (i.e. long-COVID) in patients after mild COVID-19 is debated. Within a cohort of healthcare workers (HCW), frequency and risk factors for symptoms compatible with long-COVID are assessed.

**Methods:** Participants answered baseline (August/September 2020) and weekly questionnaires on SARS-CoV-2 nasopharyngeal swab (NPS) results and acute disease symptoms. In January 2021, SARS-CoV-2 serology was performed; in March, symptoms compatible with long-COVID (including psychometric scores) were asked and compared between HCW with positive NPS, seropositive HCW without positive NPS (presumable a-/pauci-symptomatic infections), and negative controls. Also, the effect of time since diagnosis and quantitative anti-S was evaluated. Poisson regression was used to identify risk factors for symptom occurrence.

**Results:** Of 3’334 HCW (median 41 years; 80% female), 556 (17%) had a positive NPS and 228 (7%) were only seropositive. HCW with positive NPS more frequently reported ≥1 symptom compared to controls (73%*vs*.52%, p<0.001); seropositive HCW without positive NPS did not score higher than controls (58%vs.52%, p=0.13), although impaired taste/olfaction (16%*vs*.6%, p<0.001) and hair loss (17%*vs*.10%, p=0.004) were more common. Exhaustion/burnout was reported by 24% of negative controls. Many symptoms remained elevated in those diagnosed >6 months ago; anti-S titers correlated with high symptom scores. Acute viral symptoms in weekly questionnaires best predicted long-COVID symptoms. Physical activity at baseline was negatively associated with neurocognitive impairment and fatigue scores.

**Conclusions:** Seropositive HCW without positive NPS are only mildly affected by long-COVID. Exhaustion/burnout is common, even in non-infected HCW. Physical activity might be protective against neurocognitive impairment/fatigue symptoms after COVID-19.

**summary:** In this prospective healthcare worker cohort, participants with SARS-CoV-2-positive nasopharyngeal swab were most likely to report long-COVID symptoms, whereas seropositive participants without positive swab were only mildly affected. Physical activity at baseline was negatively associated with neurocognitive impairment and fatigue.

## Introduction

Emerging evidence points to different long-lasting symptoms after severe acute respiratory syndrome coronavirus-2 (SARS-CoV-2) infection, also called long-COVID [1]. The most commonly reported symptoms are fatigue, shortness of breath, chest pain, olfactory dysfunction, and neurocognitive impairment. Data suggest that long-COVID is a complex multisystem illness and that individual long-COVID symptoms are caused by different biological mechanisms that are not mutually exclusive [2-5]. Symptoms seem to decrease over the first 3 months without significant decline thereafter, and follow-up durations of up to 12months have been reported [4, 6-10]. Risk factors for long-COVID include female sex, high body mass index (BMI), and severity of COVID-19 [4, 6-8]. Most data on long-COVID come from cohorts of hospitalized patients [5]. However, the burden of long-COVID in patients with mild or even asymptomatic SARS-CoV-2 infection is largely unknown. Also, adequate control groups are often lacking, which is crucial in the light of the non-specific nature of many long-COVID symptoms [2, 11, 12].

In this prospective multicenter healthcare worker (HCW) cohort, we compared the frequency of symptoms compatible with long-COVID (including several established psychometric scores) between HCW with positive nasopharyngeal swab (NPS), HCW with presumably a- or pauci-symptomatic (i.e. only seropositive) COVID-19, and negative controls. Furthermore, the effect of time since diagnosis and the concentration of antibodies against the viral spike protein was assessed. We also sought to identify potentially modifiable factors associated with the occurrence of these symptoms.

## Methods

### Study design, population and procedures

This cohort was initiated in late summer 2020, after the first COVID-19 wave in Switzerland (Figure S1). Hospital employees from 23 healthcare institutions located in Northern and Eastern Switzerland were eligible; study enrolment took place in July and August 2020 [13]. The study was approved by the ethics committee of Eastern Switzerland (#2020-00502). Participants answered a baseline questionnaire upon inclusion and were then prospectively followed. They answered weekly electronic questionnaires, including symptoms compatible with viral respiratory infection, date/result of any SARS-CoV-2 NPS, and - starting from January 2021 - receipt of any COVID-19 vaccine. In January/February 2021, SARS-CoV-2 nucleocapsid antibodies were measured and participants were informed about their serology results. Finally, participants answered a questionnaire in March 2021, which asked about the presence of symptoms associated with long-COVID and contained four psychometric scores (see below). Participants vaccinated in the week before the long-COVID questionnaire were excluded.

### SARS-CoV-2 diagnostics

HCW were requested to get tested in case of compatible symptoms according to national recommendations. These included fever and/or the presence of any respiratory symptom (i.e. shortness of breath, cough, or sore throat). Detection of SARS-CoV-2 was made by polymerase chain reaction (PCR) or rapid antigen test (RAT), depending on the method used in the participating institutions. Self-reported swab results were validated as described previously [14].

Anti-nucleocapsid protein antibodies (anti-N) were determined to assess seropositivity against SARS-CoV-2, as described previously [13]. For most of anti-N positive HCW, anti-spike protein antibodies (anti-S) were measured.

### Predictor variables and outcomes

Our main predictor variable was SARS-CoV-2 positivity, which was further subdivided into HCW with positive self-reported NPS irrespective of serology result (suggesting symptomatic infection), and those with positive anti-N AND either negative or no NPS result (suggesting a-/pauci-symptomatic infection). SARS-CoV-2 negative HCW (i.e. seronegative without positive NPS) served as control group. As main outcome, we calculated a sum score of self-reported symptoms (among cough, dyspnea, headache, muscle/limb pain, fever/fevery feeling, chills, impaired olfaction, chest pain, heart palpitations, dizziness, exhaustion, and tiredness). Individual symptoms were also analyzed. Furthermore, we calculated the sum score of four well-established psychometric measurements, including the 16-item Rivermead Post-Concussion Questionnaire score (RMEAD) as a measure for neurocognitive impairment, the 9-item Fatigue Severity Scale (FSS), the 8-item Patient Health Questionnaire (PHQ) (without the question on sleep quality) for depression, and the 7-item General Anxiety Disorder (GAD) score [15, 16] (Table S1). Because of the considerable number of missing values for the psychometric scores, a complete case analysis was also performed.

### Influence of time since diagnosis and anti-S titers

For HCW with SARS-CoV-2-positive NPS, we further determined the influence of the time elapsed since diagnosis on all outcomes. HCW were therefore classified into the following four groups, according to the time of positive swab: 0 to 4 weeks (‘acute infection’); 5 to 12 weeks (‘acute post-COVID’); 13 to 24 weeks (‘long post-COVID’); and >24 weeks (‘persistent post-COVID’) prior to the questionnaire [17]. Again, SARS-CoV-2 negative HCW served as control group.

Similarly, for those with positive NPS and available anti-S results, we determined the influence of the anti-S titer on the symptom sum score and the psychometric measurements. HCW were classified into four groups, according to the quartile of their anti-S titer. Because this analysis was adjusted for time since positive NPS, we did not include a negative control group.

### Statistical analyses

The proportion of HCW reporting any and reporting individual long-COVID symptoms was related to SARS-CoV-2 status (NPS-positive/only seropositive/negative) with Chi-squared tests and the calculation of odds ratios (OR), and to time interval since positive swab with Chi-squared tests for trend in proportions.

The symptom sum score and scores of the psychometric measurements were analyzed using univariable quasi-Poisson regression to estimate mean scores per patient group. Quasi-Poisson models include an overdispersion parameter to account for increased error variation. For the association between anti-S and each of the scores, we used a generalized additive model to adjust for time since positive NPS (included as a smooth term). Multivariable quasi-Poisson regression was used to assess the independent association of SARS-CoV-2 positivity with symptom scores. Potential confounders included baseline health data, social determinants, job details, cultural and physical activities at baseline, and receipt of any COVID-19 vaccine. As a measure of disease severity, the total number of acute viral symptoms reported during the four follow-up weeks with the highest number of symptoms was included as co-variable. For definitions and further details including variable selection for multivariable analysis and handling of missing data see supplements.

## Results

### Study population

We included 3’346 participants, representing 20% of the eligible population of the participating hospitals. Twelve HCW with recent SARS-CoV-2 diagnosis (within 4 weeks before the long-COVID questionnaire) were only included for the analysis on symptom frequency according to time elapsed since diagnosis, but not for the other analyses (Figure 1). For the remaining 3’334 HCW, median age was 40.5 years, 80% were female; 47% were nurses and 16% physicians; patient contact was reported by 83%. These characteristics were similar to the entire eligible population [13]. Baseline characteristics by SARS-CoV-2 positivity are shown in Table 1. COVID-19 was diagnosed in 784 (24%) HCW, either by positive NPS and any serology result (n=556, 71%), or by positive serology only (n=228, 29%); 2’550 HCW (76%) were seronegative and reported either negative or no NPS (Figure 1).

**Table 1.** Characteristics of participants with SARS-CoV-2-positive swab, seropositive participants (no positive swab), and seronegative participants (no positive swab).

| Variable | Levels | Positive NPS<br>n = 556 | Only seropositive<br>n = 228 | Negative<br>n = 2550 |
| --- | --- | --- | --- | --- |
| **Anthropometrics** |  |  |  |  |
| Age (years) | median (range) | 38.9 (16.8-63.5) | 37.9 (17.1-63.9) | 41.0 (16.5-72.6) |
| Sex | male | 98 (18%) | 45 (20%) | 529 (21%) |
|  | female | 453 (81%) | 182 (80%) | 2003 (79%) |
| Body mass index (kg/m<sup>2</sup>) | median (range) | 23.5 (16.5-47.3) | 23.8 (17.7-40.6) | 23.3 (14.3-50.0) |
| Pregnancy (females) | yes | 24 (5%) | 9 (5%) | 67 (3%) |
|  | no | 386 (85%) | 158 (87%) | 1825 (91%) |
| **Social determinants (at baseline)** |  |  |  |  |
| Ethnicity | Caucasian | 550 (99%) | 225 (99%) | 2496 (98%) |
|  | other | 6 (1%) | 3 (1%) | 54 (2%) |
| Children ≤ 6 years in household | yes | 97 (17%) | 34 (15%) | 429 (17%) |
|  | no | 459 (83%) | 194 (85%) | 2121 (83%) |
| Social leisure activities | sports club, yoga, gym | 240 (43%) | 103 (45%) | 1141 (45%) |
|  | only other(s) | 178 (32%) | 68 (30%) | 810 (32%) |
|  | none | 138 (25%) | 57 (25%) | 599 (23%) |
| **Risk profile (at baseline)** |  |  |  |  |
| Comorbidity | yes (any disease) | 199 (36%) | 78 (34%) | 929 (36%) |
| Medication intake | yes (any drug) | 262 (47%) | 111 (49%) | 1162 (46%) |
| Active smoking | yes | 61 (11%) | 25 (11%) | 413 (16%) |
|  | no | 495 (89%) | 203 (89%) | 2137 (84%) |
| Alcohol intake | > 1 drink /week | 219 (39%) | 85 (37%) | 1172 (46%) |
|  | ≤ 1 drink /week | 337 (61%) | 143 (63%) | 1378 (54%) |
| **Work-related factors (at baseline)** |  |  |  |  |
| Profession | nurse | 329 (59%) | 131 (57%) | 1033 (41%) |
|  | physician | 70 (13%) | 25 (11%) | 428 (17%) |
|  | other | 141 (25%) | 59 (26%) | 980 (38%) |
| Patient contact | yes | 482 (87%) | 183 (80%) | 1924 (75%) |
|  | no | 52 (9%) | 25 (11%) | 473 (19%) |
| Contacts with COVID-19 patients | >20 | 227 (41%) | 103 (45%) | 637 (25%) |
|  | 1–20 | 177 (32%) | 58 (25%) | 869 (34%) |
|  | none | 79 (14%) | 40 (18%) | 806 (32%) |
| Working in intensive care | yes | 40 (7%) | 17 (7%) | 187 (7%) |
|  | no | 516 (93%) | 211 (93%) | 2363 (93%) |
| Work load >80% | yes | 304 (55%) | 140 (61%) | 1329 (52%) |
|  | no | 252 (45%) | 88 (39%) | 1221 (48%) |
| **COVID-19 history** |  |  |  |  |
| Serology | positive | 453 (81%) | 228 (100%) | 0 |
|  | negative | 56 (10%) | 0 | 2550 (100%) |
| Nasopharyngeal swab(s) | any positive | 556 (100%) | 0 | 0 |
|  | all negative | 0 | 113 (50%) | 1475 (58%) |
|  | none reported | 0 | 115 (50%) | 1075 (42%) |
| Mean no symptoms /week | median (range) | 0.3 (0.0-4.2) | 0.1 (0.0-5.0) | 0.1 (0.0-4.0) |
| SARS-CoV-2 vaccine | yes | 70 (13%) | 75 (33%) | 1123 (44%) |
|  | no | 486 (87%) | 153 (67%) | 1427 (56%) |

NPS, nasopharyngeal swab

**Figure 1.**
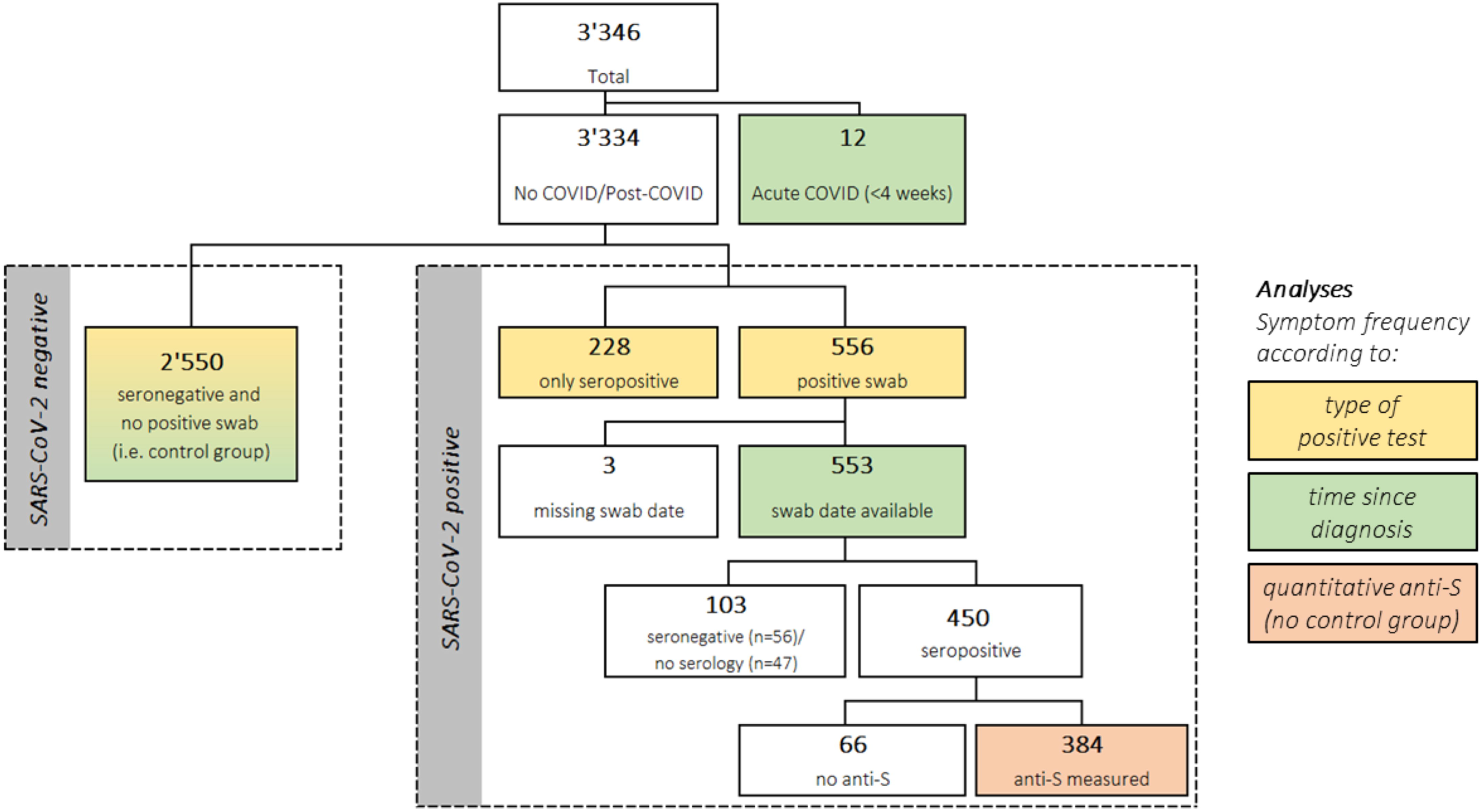
Study flow showing the total population and the sub-populations analyzed for specific analyses.

### Frequency of long-COVID-19 symptoms

The proportion of HCW reporting ≥1 symptom was higher for HCW with positive NPS compared to negative controls (73% vs. 52%, p<0.001), but not for seropositive HCW without positive NPS (58% vs. 52%, p=0.13). The most common symptoms were exhaustion/burnout (33% in NPS-positive *vs*. 25% in only seropositive *vs*. 24% in negative controls) and weakness/tiredness (34% *vs*. 25% *vs*. 22%). Impaired taste/olfaction (33% *vs*. 16% *vs*. 6%) and hair loss (17% *vs*. 17% *vs*. 10%) were the only symptoms which were significantly more common in only seropositive HCW compared to negative controls. Symptoms of acute infection such as fever, chills, headache or dyspnea, were not more common in SARS-CoV-2 positive participants (Figure 2, Table S2).

**Figure 2.**
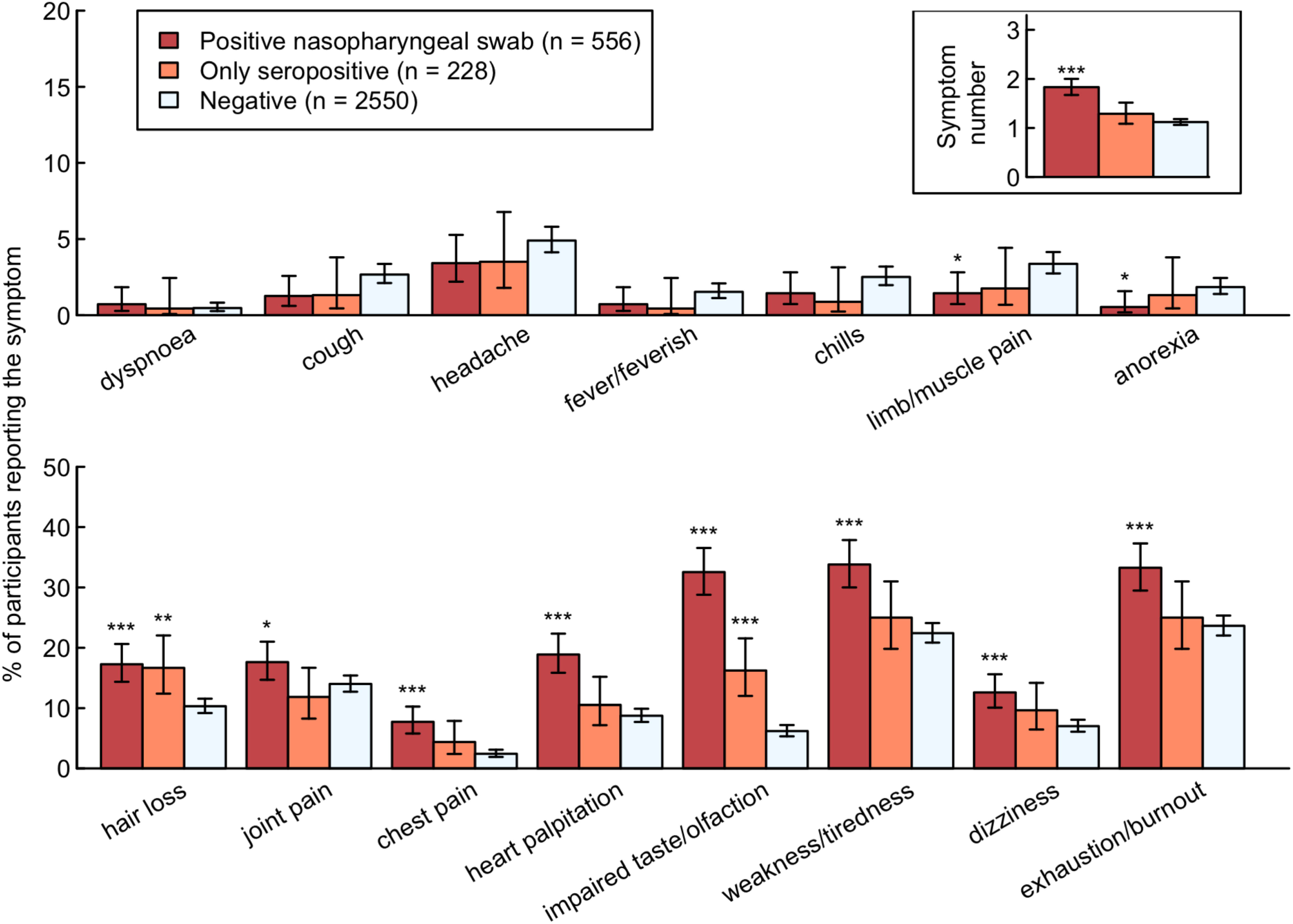
Percentage of healthcare workers (HCW) reporting symptom and total number of reported symptoms (upper right corner) by HCW with positive SARS-CoV-2 swab (red), seropositive HCW without positive swab (orange), and seronegative HCW without positive swab (light blue). Statistics: Positive HCW are compared to negative HCW, respectively (*** p-value <0.001; ** p-value 0.001-0.01; * p-value 0.01-0.05).

Similarly, NPS-positive participants (but not seropositive participants without positive swab) showed significantly higher scores in the RMEAD, FSS and PHQ compared to negative controls, whereas this was not the case for the anxiety (GAD) score (Figure 3). The complete case analysis showed similar results (Figure S2). A fatigue score of 36 or more, indicating clinically relevant fatigue, was obtained for 260 (10.6%) of SARS-CoV-2 negative participants, for 22 (10.0%) of seropositive participants without positive swab (OR 0.9, 95% CI 0.6-1.5, p=0.86) and for 130 (23.7%) of NPS-positive participants (OR 2.6, 95% CI 2.1-3.3, p<0.001).

**Figure 3.**
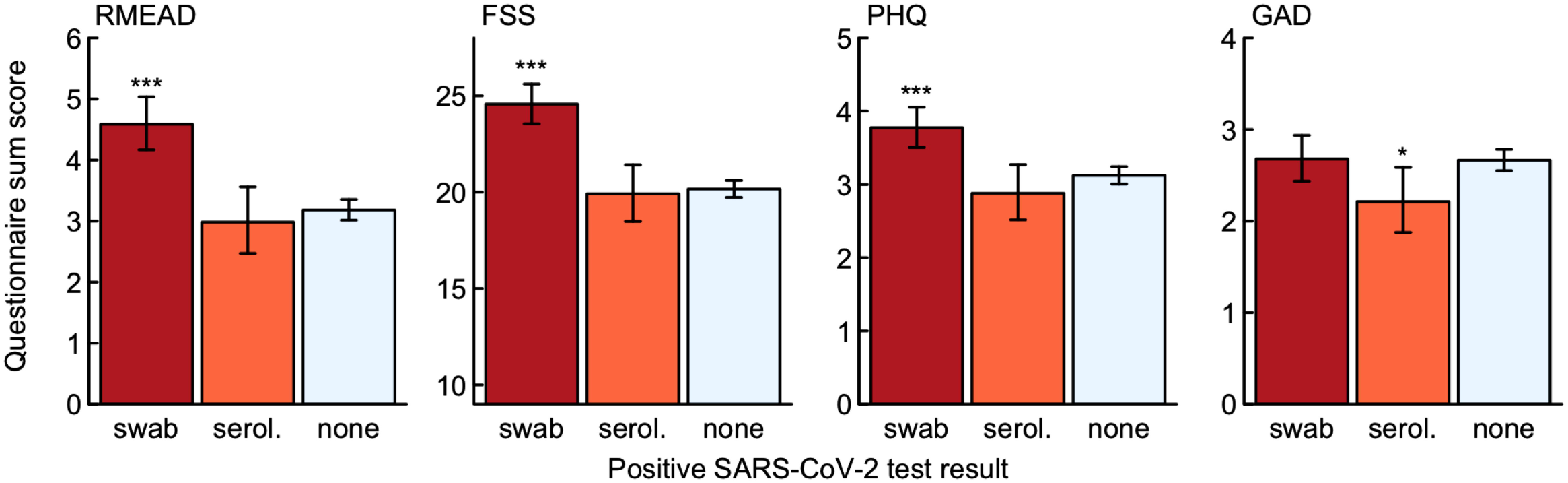
Sum score for the post-concussion (RMEAD), fatigue severity (FSS), depression (PHQ), and anxiety (GAD) scores by healthcare workers (HCW) with positive SARS-CoV-2 swab (red), seropositive HCW without positive swab (orange), and seronegative HCW without positive swab (light blue). Statistics: Positive HCW are compared to negative HCW, respectively (*** p-value <0.001; ** p-value 0.001-0.01; * p-value 0.01-0.05).

### Time since diagnosis

For this analysis, we included 565 participants with positive NPS after a median of 117 days (interquartile range 93-147) after infection and 2’550 SARS-CoV-2 negative controls (Figure 1). The total number of symptoms declined after acute infection (p<0.001), but was still higher after 24 weeks compared to non-infected controls (Figure 4). Cough, dyspnea, headache, chills, limb/muscle pain, and anorexia were mainly reported within 4 weeks after acute infection, but rarely thereafter. Other symptoms such as impaired taste and weakness/tiredness were frequent during acute infection and remained significantly more common after 24 weeks compared to the control; hair loss was not reported within 4 weeks after acute infection, but only in the post-acute phase.

**Figure 4.**
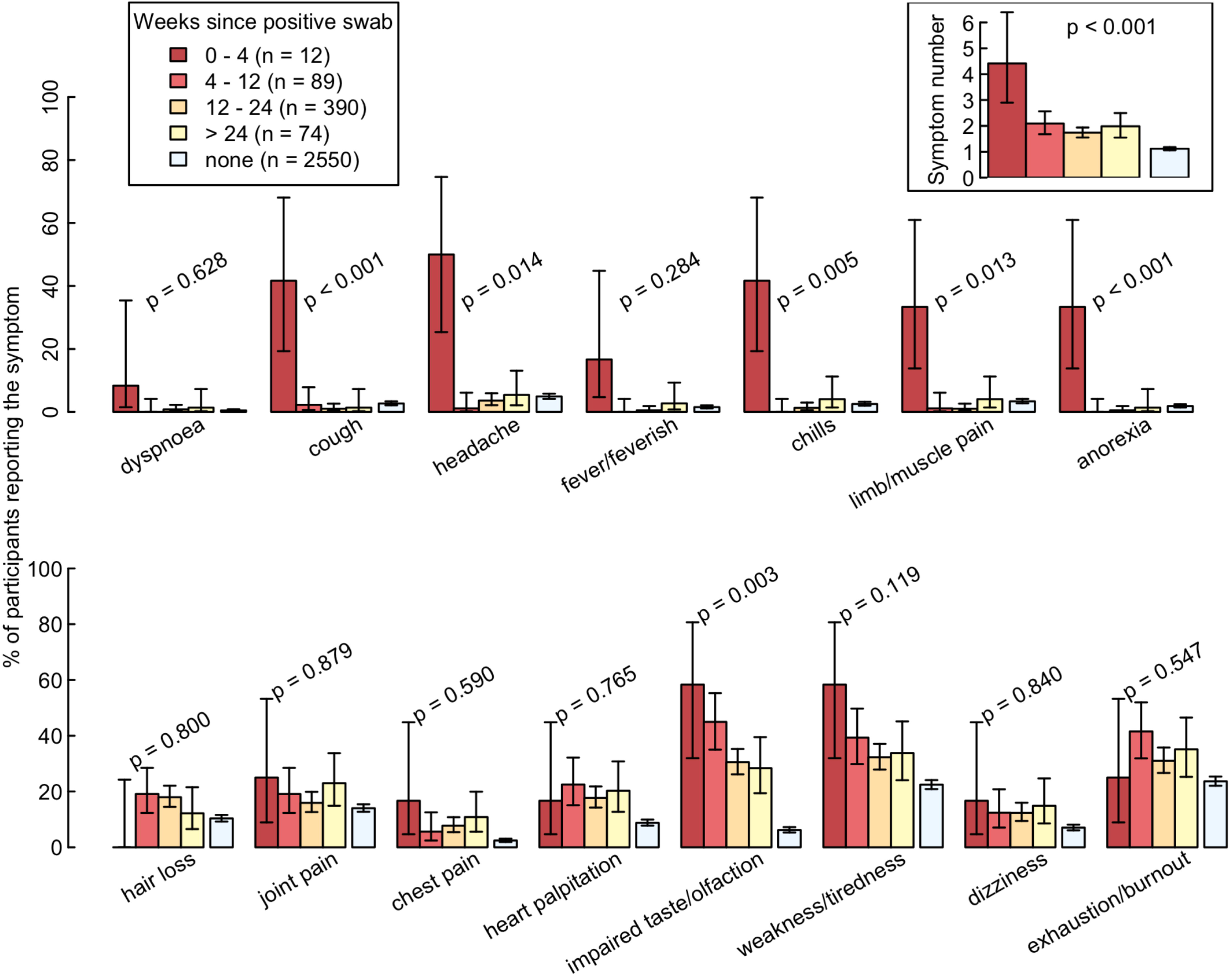
Percentage of healthcare workers reporting individual symptoms and total number of reported symptoms (upper right corner) according to weeks since positive SARS-CoV-2 swab, compared to negative controls. Statistics: P-values from tests for trends over the four categories, without negative controls.

For the psychometric measurements, there was no correlation between sum scores and time since positive swab, such that scores remained significantly higher in the RMEAD, FSS, and PHQ for those with an infection history more than 24 weeks ago (Figure S3).

### Antibody concentration against the viral spike protein (anti-S titers)

We included 384 participants with positive NPS and available anti-S results for this analysis (Figure 1). Adjusted for time since positive swab, the quartile with the highest anti-S titers consistently scored highest for symptom numbers and all psychometric measurements. However, statistical significance was only reached for the FSS when evaluating the trend across all quartiles (Figure 5).

**Figure 5.**
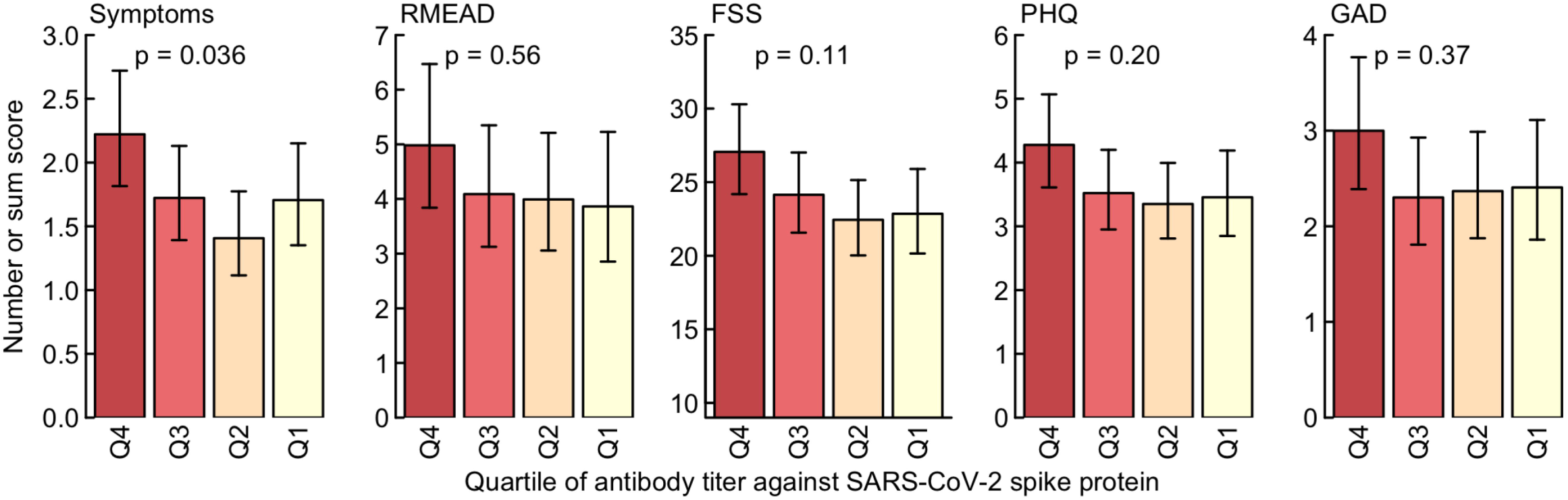
Sum score for symptoms, the post-concussion (RMEAD), fatigue severity (FSS), depression (PHQ), and anxiety (GAD) scores according to quartile of SARS-CoV-2 anti-spike titers. Results are adjusted for time since positive swab. Statistics: P-values from tests for linear trends over the four categories.

### Multivariable analyses

Results of univariable analysis regarding sum score of the long-COVID questions is shown in Table S3. In multivariable analysis, SARS-CoV-2 positivity, younger age, female sex, increased BMI, any comorbidity and any medication at baseline as well as working as a nurse were significantly associated with the symptom sum score. The strongest risk factor was the number of acute viral symptoms reported in the weekly questionnaires; COVID-19 vaccination (mostly administered *after* SARS-CoV-2 infection, where applicable) was not associated with the outcome (Figure 6). Multivariable analyses for the RMEAD and FSS score showed similar results. However, SARS-CoV-2 positivity was not associated with the RMEAD score after accounting for all potential confounders. Having children <6 years in the same household and physical activities at baseline were negatively associated with both scores, while working in intensive care was positively associated with the FSS score (Figure S4).

**Figure 6.**
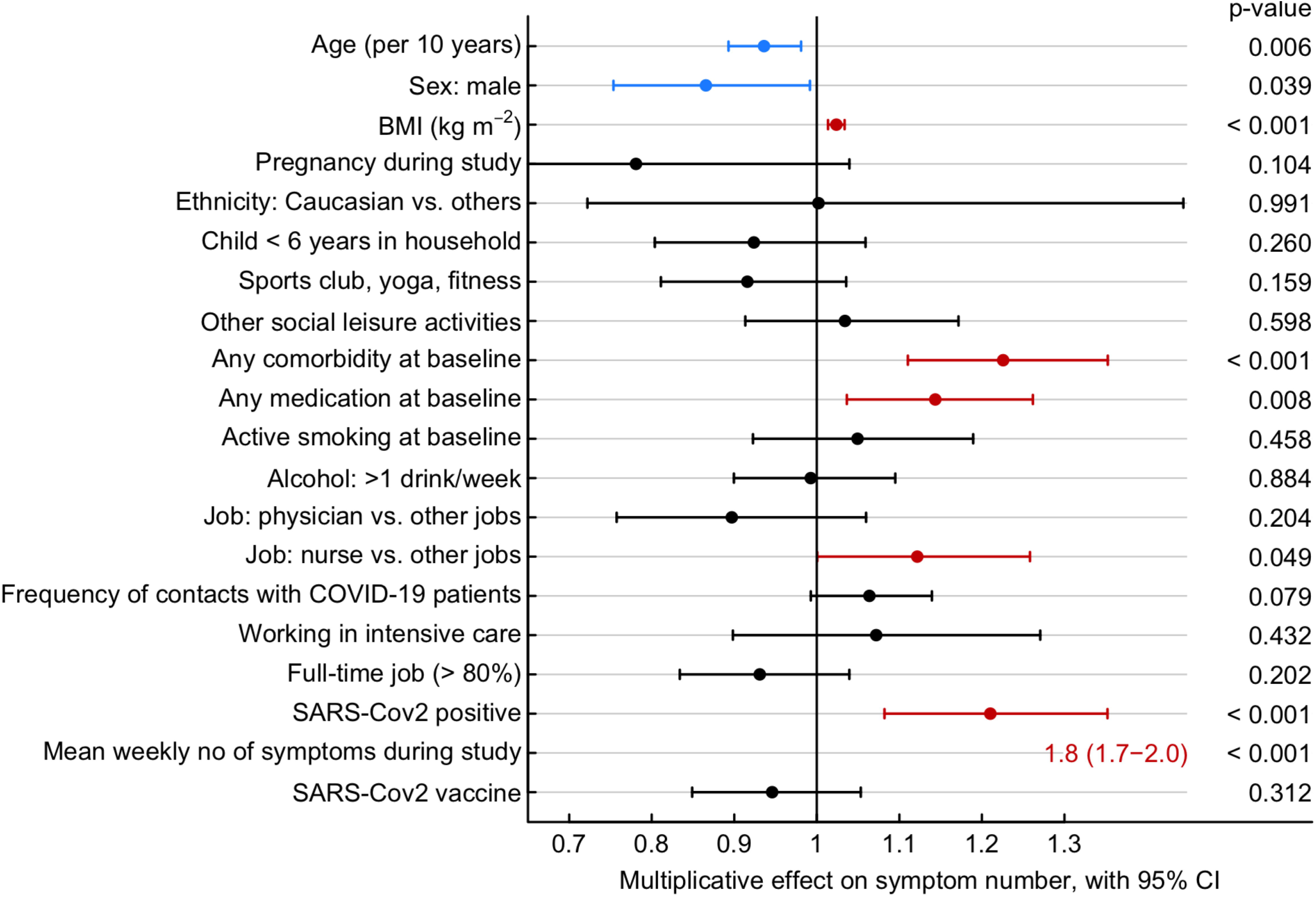
Forest plot showing the multiplicative effect of potential influential factors on the number of long-COVID symptoms, as determined through multivariable Poisson regression. Factors in blue are negatively, those in red positively associated with the number of symptoms.

## Discussion

In this prospective cohort of SARS-CoV-2 infected and non-infected HCW, symptoms compatible with long-COVID were most common in HCW with a positive NPS, whereas seropositive HCW without positive NPS were only mildly affected. For most symptoms, we did not observe a significant attenuation with increasing time since diagnosis. Physical activity at baseline showed a negative association with neurocognitive impairment and fatigue symptoms. The main strengths of our study are the inclusion of a non-infected control group, the prospective follow-up with weekly questionnaires capturing acute viral symptoms, and the large number of serologies performed, which allowed us to identify HCW with asymptomatic seroconversion and to ascertain truly negative controls with high reliability.

The main finding of our study is that seropositive participants without positive swab, representing presumable a- or pauci-symptomatic disease, showed similar symptom frequency as the negative control group, both for the symptom sum score as well as for the psychometric scores. This is in line with a study evaluating the physical endurance of young recruits after a COVID-19 outbreak, where those with asymptomatic infection – in contrast to symptomatic recruits – did not show reduced maximal aerobic capacities [18]. In an Italian cohort assessing post-COVID symptoms at 6 months after infection, 40% of those with mild disease reported post-COVID symptoms (similar to those with severe disease), whereas only 5% of asymptomatic participants reported post-COVID symptoms [19].

Impaired taste/olfaction and hair loss were the only symptoms being more prevalent in seropositive participants without positive swab compared to negative controls. Impaired taste/olfaction was also the only symptom showing a significant attenuation over time, but was still frequently reported by those infected >24 weeks ago. This is in accordance with multiple other reports, which classifies impaired taste/olfaction as cardinal symptom for long-COVID. Of note, loss of taste has been associated with a lower quality of life and increased risk for depression [20, 21], which underlines the pathological significance of this symptom. A possibly under-recognized symptom of long-COVID is hair loss, which has in fact been reported as one of the most common symptoms in some studies, with more than 25% of patients being affected [22, 23]. In a case series of almost 200 patients from Spain, a prior history of fever was the only factor associated with hair loss after SARS-CoV-2 infection [24]. In our study, hair loss was reported by 17% of positive participants.

The symptoms most commonly reported were weakness and exhaustion. This is in line with other reports, showing that up to 55% of COVID-19 patients can be affected by these long-term sequelae [15]. However, these symptoms were also reported by almost one fourth of non-infected controls. This shows that including a control group to assess these non-specific symptoms is of uttermost importance. Irrespective of SARS-CoV-2 infection, many HCW are exhausted one year into the pandemic, which is also reflected in the results of the multivariable analysis for the fatigue score, where working in intensive care was identified as independent risk factor. Several other studies have reported similar findings [11, 25, 26].

The number of acute viral symptoms was the strongest risk factor for long-COVID symptoms, as shown previously [6]. In line with this finding, anti-S concentrations, which is a proxy for the severity of acute disease [27], was associated with the presence of long-COVID symptoms. Similar results have been reported previously [12, 19]. These results are plausible and anti-S titers could therefore be used to identify high-risk populations, which could particularly benefit from early intervention therapies. However, acute viral symptoms were associated with long-COVID independent of documented SARS-CoV-2 infection, which can be explained by the fact that probably not all symptomatic HCW underwent testing for SARS-CoV-2; at the same time, the sensitivity of the serology test used in our study is only about 90% [28]. Other viral diseases have also been postulated to cause either long-term respiratory symptoms in the case of respiratory syncytial virus [29],chronic fatigue after infection with influenza or Epstein Barr virus [30, 31] or long-COVID clinical features after influenza [32, 33]. However, we do not know in how far other infections might have played a role in our cohort.

Other risk factors, such as female sex, increased BMI, or baseline comorbidities have also been identified by others as risk factors for long-COVID [9]. For age, data are conflicting. Some studies have reported more long-COVID symptoms with increasing age [6], whereas others have reported an association with younger age, being in line with our data [34]. Of note, SARS-CoV-2 vaccination (administered *after* SARS-CoV-2 infection in the large majority of our patients) did not reduce the number of long-COVID symptoms, as has been suggested in a previous study [35]. However, a recent study showed that if people are vaccinated *before* SARS-CoV-2 infection, the risk for long-COVID seems to be decreased [36].

Interestingly, we found physical activity at baseline to be negatively associated with the neurocognitive impairment and fatigue scores, suggesting a protective role in the prevention of these symptoms. The facts that i) other social activities were not associated with decreased risk and that ii) ‘paced’ physical activity is also part of successful treatment bundles for long-COVID both suggest a true effect, although residual confounding is possible [37]. Further study is needed to evaluate the potential role of physical activity in the prevention and also therapy of long-COVID.

We also found having children <6 years to be negatively associated with neurocognitive impairment and fatigue, a finding which is not readily intuitive. However, increased physical activity during COVID-19 in HCW living with children has been reported, which may also explain the positive effects found in our study [38].

Our study has limitations. First, assessing the presence of long-COVID symptoms at only one time point might have led to underestimation of their prevalence as symptoms are reported to be fluctuating [5]. Second, we cannot exclude that certain individuals with COVID-19 did not show any seroconversion or that anti-N had again waned below the detection level at time of our blood draw. However, we consider this effect to be small as most individuals, even after mild/moderate infection, show robust and prolonged antibody reaction [39]; also, individuals were asked to report any positive NPS, which further reduces the risk of misclassification. Third, participants knew about their SARS-CoV-2 status (both for serology and NPS) at time of the long-COVID questionnaire, which might be a source of recall bias. However, the fact that participants with the highest concentration of anti-S (titers not known to participants) had higher symptom scores suggests a true effect. Fourth, the delta variant, which was not yet the predominant strain when the study was conducted, might differently impact the occurrence of long-COVID symptoms [40]. Fifth, we did not assess long-COVID symptoms at baseline which precludes us from estimating the true impact of the pandemic on the prevalence of these symptoms. Nevertheless, the inclusion of a non-infected control group allowed us to assess the independent impact of SARS-CoV-2 on the occurrence of these often non-specific symptoms.

In conclusion, we show that SARS-CoV-2 seropositive HCW without positive NPS are only mildly affected by long-COVID. However, if present, most symptoms including neurocognitive and fatigue symptoms persist, even for those with a diagnosis >6 months ago. The main predictor for long-COVID is the severity of acute disease, also reflected in the highest symptom frequency among those with high anti-S titers. Exhaustion and tiredness are commonly reported among HCW, also in those without SARS-CoV-2 infection. This is probably a consequence of the general burden, which the pandemic is having on this particularly exposed population. Further studies should evaluate the potential protective role of physical activity in the prevention of post-concussion and fatigue symptoms after SARS-CoV-2 infection.

## Data Availability

All data produced in the present study are available upon reasonable request to the authors

## Acknowledgment

We would like to thank the employees of the participating healthcare institutions who either took part in this study themselves or supported it. Furthermore, we thank the laboratory staff for shipment, handling and analysis of the blood samples.

---

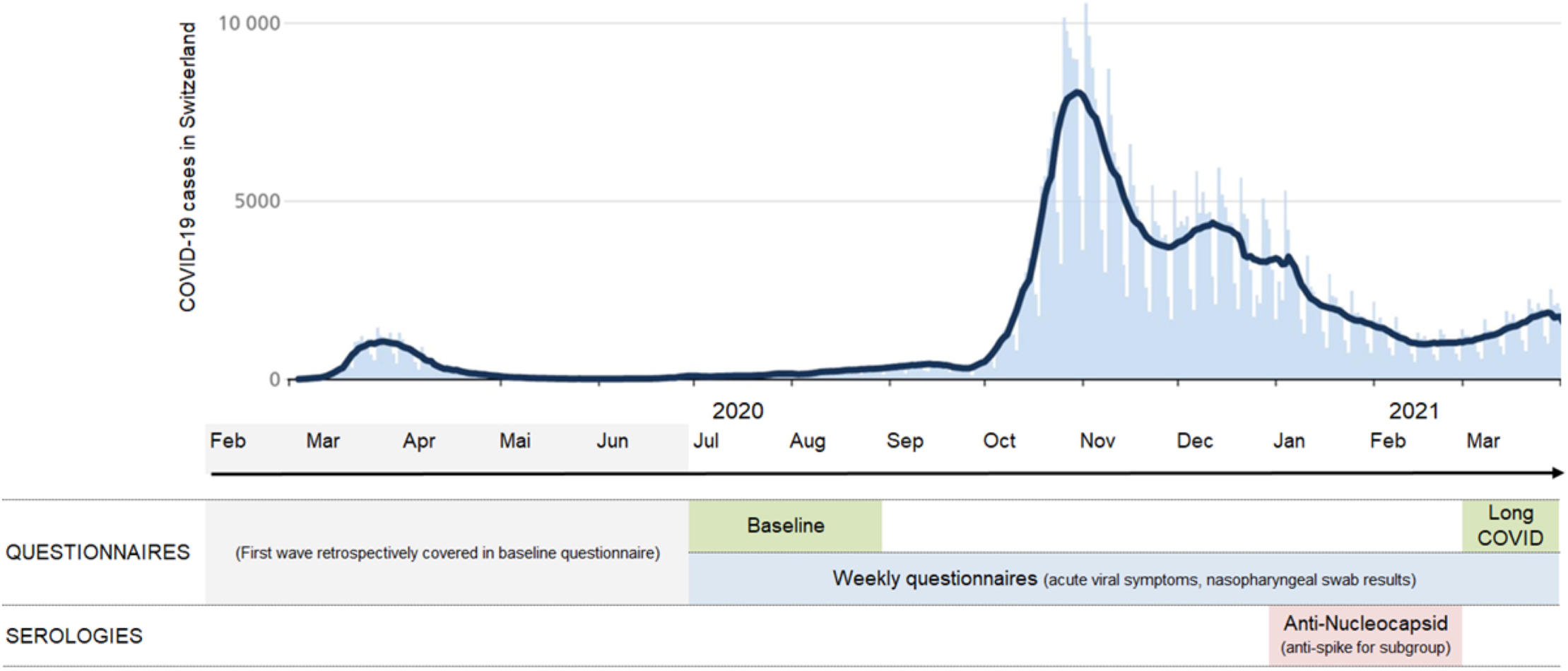

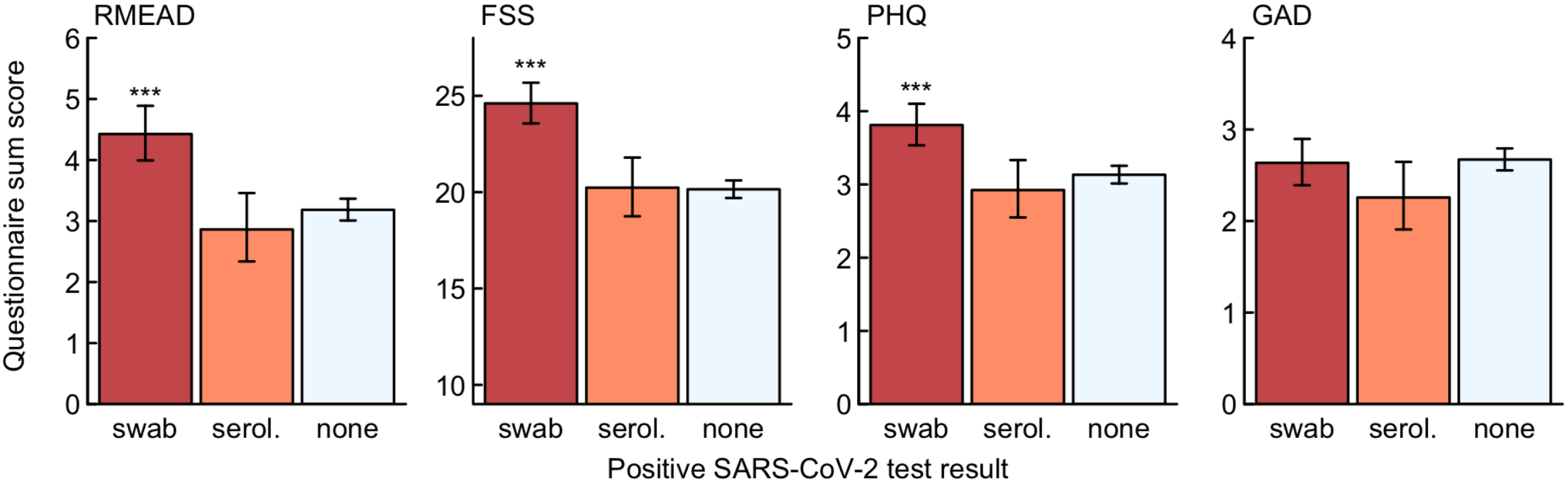

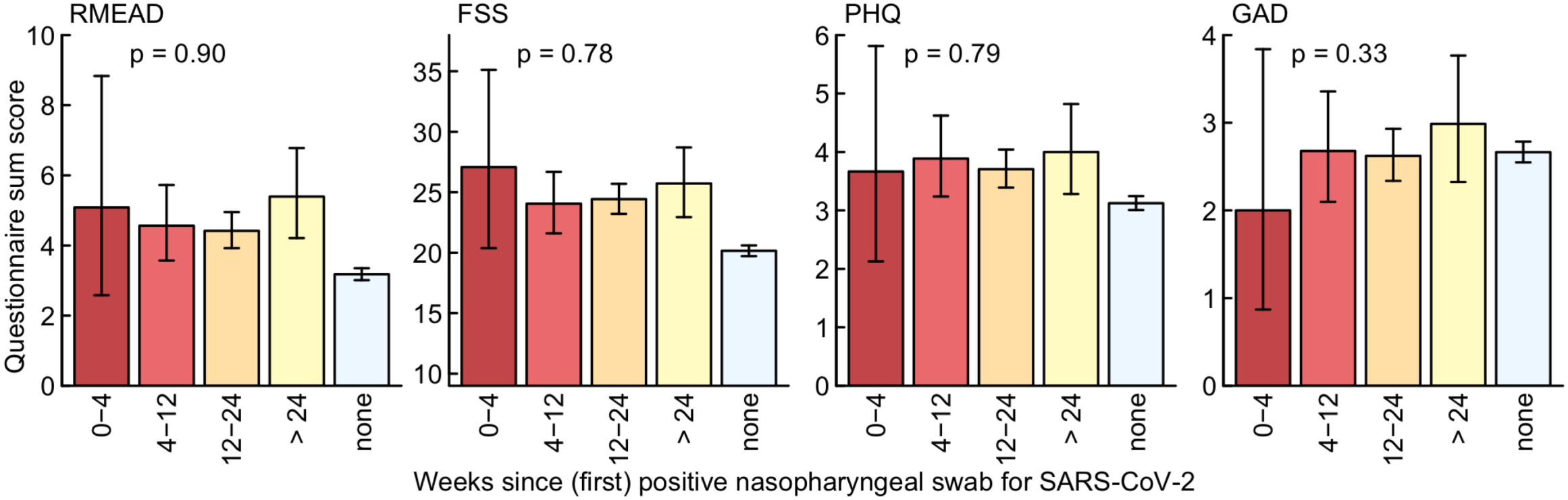

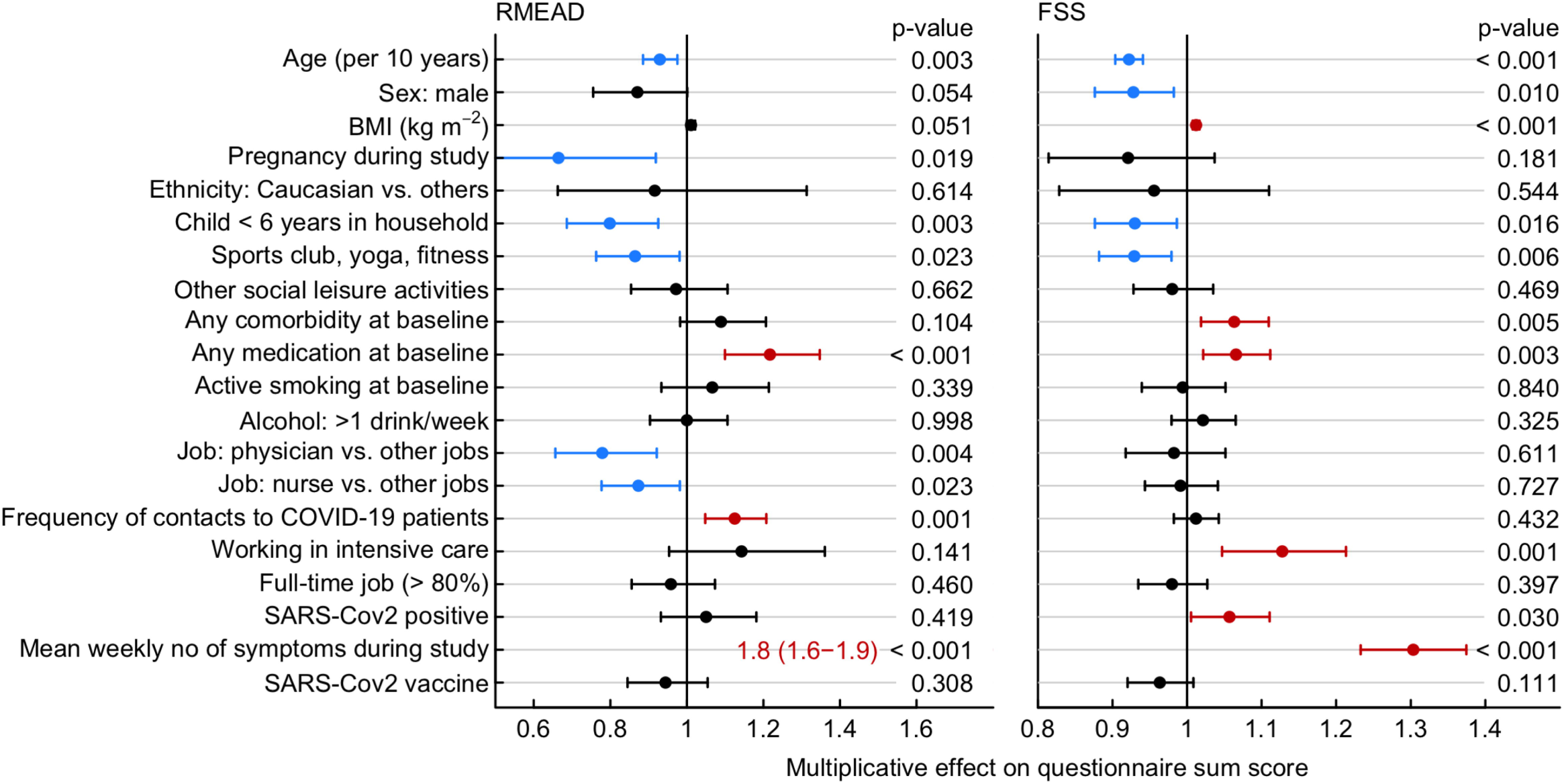

