## Supplemental_material for "Symptoms compatible with long-COVID in healthcare workers with and without SARS-CoV-2 infection – results of a prospective multicenter cohort"

Strahm et al.

**METHODS (Supp.)**

*Psychometric scores and missing data*

Psychometric scores included the RMEAD, FSS, PHQ and GAD, consisting of 16, 9, 8 and 7 items, respectively, rated on an ordinal scale with 4 to 7 levels (Table S1). Scores of the RMEAD scale were transformed from the original 0−4 points scale to a 0−3 points scale by subtracting 1 point from all positive scores, to account for the fact that score 1 means "no longer a problem", i.e. currently equivalent to score 0. The main outcome for each questionnaire was a sum score calculated as the sum of individual items. For the FSS, the sum score was dichotomized as <36 (mean score <4, no clinically relevant fatigue) and ≥36 (mean score ≥4, clinically relevant fatigue).

For HCW skipping items of the neuropsychological scores, missing value imputation was performed in order to obtain valid sum scores provided that no more than 20% of a questionnaire's items had been skipped. Participants who skipped more than 20% of a questionnaire's items were ignored when fitting the models used for imputation and no sum score was calculated. Missing value imputation was performed through multivariate imputation by chained equations (R package mice, version: 3.13.0) using predictive mean matching and 10 iterations over the individual items. Three separate series of imputations were performed for each questionnaire in order to evaluate the robustness of the sum scores obtained from a single series of imputations.

*Statistical analyses*

The number of symptoms was analyzed using Poisson regression with overdispersion parameter (“quasipoisson” distribution family). Mean symptom numbers and sum scores with 95% confidence intervals were derived from the coefficients of Poisson models without intercept.

Trends across the four time intervals since positive swab were tested using linear contrasts after fitting a model with the four groups. To correct for the fact that anti-spike titers depend on the time since diagnosis, the association of sum scores with anti-spike titers was derived from generalized additive models including a smooth term for time since positive swab.

Multivariable Poisson models were used to analyze the effect of risk factors on the number of long-COVID symptoms. Risk factors were selected for multivariable analysis based on expected importance and on the number of missing values. We used uni- and multivariable linear regression analysis to calculate the (independent) multiplicative effect of predictor and confounder variables on the symptom sum score. Similarly, multivariable analysis was also performed on the RMEAD and the FSS score, but not on the PHQ-8 (rationale: one question missing) and the GAD-9 score (rationale: no effect in univariable analysis).

**TABLES (Supp.)**

Table S1. Items and answer options of psychometric scores used (RMEAD, FSS, PHQ and GAD) as well as % of participants with problem among SARS-CoV-2 negative and SARS-CoV-2 positive participants (significant results in red).

| **Questionnaire/Question** | **Answer options** | **% with problem** | | **Odds ratio** | **p-value** |
| --- | --- | --- | --- | --- | --- |
|  |  | negative | positive |  |  |
| **The Rivermead Post-Concussion Symptoms Questionnaire (RMEAD)** *(King, N. et al. J. Neurology 242: 587-592)* | | | | | |
| Compared to before the pandemic, do you currently suffer from | |  |  |  |  |
| Headaches | 0 = Not experienced at all  1 = No more of a problem  2 = A mild problem  3 = A moderate problem  4 = A severe problem | 25 | 27 | 1.13 | 0.201 |
| Feelings of Dizziness |  | 10 | 15 | 1.56 | <0.001 |
| Nausea and/or Vomiting |  | 4 | 7 | 1.68 | 0.003 |
| Noise Sensitivity, easily upset by loud noise |  | 9 | 12 | 1.32 | 0.037 |
| Sleep Disturbance |  | 28 | 28 | 1.03 | 0.746 |
| Fatigue, tiring more easily |  | 39 | 51 | 1.61 | <0.001 |
| Being Irritable, easily angered |  | 28 | 27 | 0.97 | 0.781 |
| Feeling Depressed or Tearful |  | 19 | 20 | 1.07 | 0.528 |
| Feeling Frustrated or Impatient |  | 29 | 24 | 0.79 | 0.014 |
| Forgetfulness, poor memory |  | 15 | 30 | 2.37 | <0.001 |
| Poor Concentration |  | 17 | 28 | 1.82 | <0.001 |
| Taking Longer to Think |  | 9 | 16 | 1.96 | <0.001 |
| Blurred Vision |  | 11 | 15 | 1.45 | 0.003 |
| Light Sensitivity, Easily upset by bright light |  | 6 | 7 | 1.28 | 0.138 |
| Double Vision |  | 1 | 1 | 1.39 | 0.393 |
| Restlessness |  | 11 | 9 | 0.8 | 0.123 |
| **Fatigue Severity Scale (FSS, English version)** *(Krupp et al, Arch Neurol 1989)* | | | | | |
| My motivation is lower when I am fatigued. | Patients are instructed to choose a number from 1-7 that indicates their degree of agreement with statement (1 indicates strongly disagree; 7, strongly agree*)* | 77 | 78 | 1.04 | 0.693 |
| Exercise brings on my fatigue. |  | 57 | 67 | 1.55 | <0.001 |
| I am easily fatigued. |  | 55 | 65 | 1.51 | <0.001 |
| Fatigue interferes with my physic al functioning. |  | 60 | 65 | 1.25 | 0.009 |
| Fatigue causes frequent problems for me. |  | 48 | 55 | 1.33 | 0.001 |
| My fatigue prevents sustained physic al functioning. |  | 39 | 50 | 1.55 | <0.001 |
| Fatigue interferes with carrying out certain duties and responsibilities. |  | 33 | 37 | 1.19 | 0.049 |
| **Patient Health Questionnaire (PHQ-9)** |  |  |  |  |  |
| Over the last 2 weeks, how often have you been bothered by any of the following problems? | | | | | |
| Little interest or pleasure in doing things | Not at all - 0 Several days - 1 More than half the days - 2  Nearly every day – 3 | Not asked | | | |
| Feeling down, depressed, or hopeless |  | 52 | 57 | 1.2 | 0.028 |
| Trouble falling or staying asleep, or sleeping too much |  | 38 | 39 | 1.04 | 0.67 |
| Feeling tired or having little energy |  | 67 | 73 | 1.34 | 0.001 |
| Poor appetite or overeating |  | 40 | 46 | 1.27 | 0.004 |
| Feeling bad about yourself — or that you are a failure or have let yourself or your family down |  | 25 | 26 | 1.05 | 0.635 |
| Trouble concentrating on things, such as reading the newspaper or watching television |  | 33 | 37 | 1.22 | 0.026 |
| Moving or speaking so slowly that other people could have noticed? Or the opposite — being so fidgety or restless that you have been moving around a lot more than usual |  | 9 | 12 | 1.44 | 0.006 |
| Thoughts that you would be better off dead or of hurting yourself in some way |  | 4 | 4 | 0.94 | 0.83 |
| **Generalized Anxiety Disorder 7** (**GAD-7)** |  |  |  |  |  |
| Feeling nervous, anxious or on edge | Not at all - 0 Several days - 1 More than half the days - 2  Nearly every day – 3 | 41 | 37 | 0.84 | 0.047 |
| Not being able to stop or control worrying |  | 21 | 20 | 0.92 | 0.445 |
| Worrying too much about different things |  | 37 | 36 | 0.94 | 0.493 |
| Trouble relaxing |  | 50 | 46 | 0.86 | 0.076 |
| Being so restless that it is hard to sit still |  | 17 | 17 | 0.99 | 1 |
| Becoming easily annoyed or irritable |  | 48 | 48 | 1.01 | 0.934 |
| Feeling afraid as if something awful might happen |  | 17 | 17 | 0.97 | 0.869 |

Table S2. Number of total symptoms and frequency (in %) of individual symptoms according to SARS-CoV-2 status (i.e. negative *vs.* only seropositive *vs.* positive swab); odds ratios (OR) and corresponding p-values comparing categories of positive participants with negative control group are derived from Fishers exact tests.

|  |  | Frequency of symptoms in each group | | | | Comparison of positive with negative participants | | | | | |
| --- | --- | --- | --- | --- | --- | --- | --- | --- | --- | --- | --- |
|  |  | no positive | only | positive | any positive | only seropositive | | positive NPS | | any positive test | |
|  | n | test result | seropositive | NPS | test result | OR | p-value | OR | p-value | OR | p-value |
| Number of symptoms^1^ |  | 1 (0-2) | 1 (0-2) | 1 (0-3) | 1 (0-3) | 1.06 | 0.131 | 1.25 | <0.001 | 1.2 | <0.001 |
| At least one symptom | 1859 | 52.0% | 57.5% | 72.5% | 68.1% | 1.25 | 0.128 | 2.43 | <0.001 | 1.97 | <0.001 |
| Dyspnoea | 17 | 0.5% | 0.4% | 0.7% | 0.6% | 0.93 | 1 | 1.53 | 0.678 | 1.36 | 0.773 |
| Cough | 78 | 2.7% | 1.3% | 1.3% | 1.3% | 0.49 | 0.308 | 0.47 | 0.071 | 0.47 | 0.034 |
| Headache | 152 | 4.9% | 3.5% | 3.4% | 3.4% | 0.71 | 0.434 | 0.69 | 0.162 | 0.69 | 0.107 |
| Feverish feeling / fever | 44 | 1.5% | 0.4% | 0.7% | 0.6% | 0.28 | 0.301 | 0.47 | 0.2 | 0.41 | 0.083 |
| Chills | 74 | 2.5% | 0.9% | 1.4% | 1.3% | 0.34 | 0.186 | 0.57 | 0.172 | 0.5 | 0.056 |
| Limb/muscle pain | 98 | 3.4% | 1.8% | 1.4% | 1.5% | 0.51 | 0.26 | 0.42 | 0.023 | 0.45 | 0.011 |
| Anorexia | 53 | 1.8% | 1.3% | 0.5% | 0.8% | 0.71 | 0.754 | 0.29 | 0.043 | 0.41 | 0.052 |
| Hair loss | 397 | 10.3% | 16.7% | 17.3% | 17.1% | 1.74 | 0.004 | 1.81 | <0.001 | 1.79 | <0.001 |
| Joint pain | 482 | 14.0% | 11.8% | 17.6% | 15.9% | 0.83 | 0.421 | 1.31 | 0.034 | 1.17 | 0.195 |
| Chest pain | 115 | 2.4% | 4.4% | 7.7% | 6.8% | 1.84 | 0.118 | 3.36 | <0.001 | 2.91 | <0.001 |
| Increased heart palpitation | 352 | 8.7% | 10.5% | 18.9% | 16.5% | 1.23 | 0.433 | 2.43 | <0.001 | 2.05 | <0.001 |
| Impaired taste or olfaction | 376 | 6.2% | 16.2% | 32.6% | 27.8% | 2.93 | <0.001 | 7.3 | <0.001 | 5.83 | <0.001 |
| Weakness / tiredness | 817 | 22.4% | 25.0% | 33.8% | 31.3% | 1.15 | 0.421 | 1.77 | <0.001 | 1.57 | <0.001 |
| Dizziness | 271 | 7.0% | 9.6% | 12.6% | 11.7% | 1.41 | 0.182 | 1.91 | <0.001 | 1.76 | <0.001 |
| Exhaustion / burnout | 845 | 23.6% | 25.0% | 33.3% | 30.9% | 1.08 | 0.705 | 1.61 | <0.001 | 1.44 | <0.001 |

^1^ median (interquartile range), OR per symptom

Table S3. Characteristics of participants and multiplicative effect on the number of symptoms (excluding participants with recent vaccination or with positive swab ≤ 4 weeks before, N=3’334).

|  | **Missing (*n*)** | ***n* or  median (range)** | **Multiplicative effect on no of symptoms with 95% CI** | ***p*-value** |
| --- | --- | --- | --- | --- |
| *Anthropometrics* |  |  |  |  |
| Age (years - effect per 10 years) | 6 | 40.5 (16.5-72.6) | 0.93 (0.90 to 0.97) | <0.001 |
| Sex (male/female) | 24 | 672/2638 | 0.73 (0.64 to 0.82) | <0.001 |
| BMI (kg/m^2^) | 9 | 23.4 (14.3-50.0) | 1.03 (1.02 to 1.04) | <0.001 |
| Blood type (0 vs. A/AB/B) | 820 | 1082/1432 | 1.03 (0.93 to 1.14) | 0.568 |
| Pregnancy (Y/N; females) | 193 | 100/2369 | 0.72 (0.53 to 0.95) | 0.025 |
| *Social determinants* |  |  |  |  |
| Ethnicity (Caucasian vs. other) | 0 | 3271/63 | 0.98 (0.72 to 1.39) | 0.927 |
| Monthly income (8 categories) | 1227 | 6000−9000 CHF | 0.91 (0.88 to 0.95) | <0.001 |
| Children ≤6 years in household (Y/N) | 0 | 560/2774 | 0.99 (0.88 to 1.11) | 0.875 |
| Social leisure activities at baseline | 0 |  |  |  |
| None |  | 794 | Reference |  |
| Sports club, yoga, fitness |  | 1484 | 0.84 (0.75 to 0.94) | 0.002 |
| Only other activities |  | 1056 | 1.01 (0.90 to 1.13) | 0.888 |
| *Risk profile* |  |  |  |  |
| Comorbidity (any; Y/N) | 151 | 1206/1977 | 1.36 (1.24 to 1.49) | <0.001 |
| Autoimmune disease |  | 82 | 1.75 (1.39 to 2.16) | <0.001 |
| Atopy/Asthma |  | 898 | 1.27 (1.15 to 1.39) | <0.001 |
| Cardiovascular/diabetes |  | 216 | 1.32 (1.12 to 1.54) | <0.001 |
| Immunosuppression/ oncologic |  | 72 | 1.69 (1.32 to 2.13) | <0.001 |
| Thyroid disease |  | 133 | 1.47 (1.21 to 1.77) | <0.001 |
| Other |  | 87 | 1.61 (1.28 to 2.00) | <0.001 |
| Medication | 0 | 1535/1799 | 1.30 (1.19 to 1.42) | <0.001 |
| Antidepressants |  | 70 | 1.40 (1.07 to 1.81) | 0.012 |
| Inhaled steroids |  | 80 | 1.62 (1.28 to 2.03) | <0.001 |
| Thyroid hormones |  | 156 | 1.38 (1.14 to 1.65) | <0.001 |
| Home remedies |  | 521 | 1.19 (1.06 to 1.34) | 0.003 |
| Active smoking (Y/N) | 0 | 499/2835 | 1.09 (0.97 to 1.23) | 0.152 |
| Alkohol intake (> 1 vs. ≤ 1 drink/week) | 0 | 1476/1858 | 0.86 (0.79 to 0.95) | 0.002 |
| *Work-related factors* |  |  |  |  |
| Profession | 138 |  |  |  |
| Other |  | 1180 | Reference |  |
| Physician |  | 523 | 0.87 (0.75 to 1.01) | 0.07 |
| Nurse |  | 1493 | 1.23 (1.11 to 1.36) | <0.001 |
| Patient contact at baseline (Y/N) | 195 | 2589/550 | 1.41 (1.24 to 1.62) | <0.001 |
| Contact with COVID-19 patients  (none, 1−20, >20; effect per level) | 338 | 925/1104/967 | 1.13 (1.06 to 1.20) | <0.001 |
| Working in intensive care (Y/N) | 0 | 244/3090 | 1.00 (0.84 to 1.18) | 0.992 |
| Work load >80% (Y/N) | 0 | 1773/1561 | 0.99 (0.90 to 1.08) | 0.766 |
| *COVID-19 history* |  |  |  |  |
| Seropositive (Y/N) | 47 | 681/2606 | 1.39 (1.26 to 1.54) | <0.001 |
| Positive swab (Y/N) | 0 | 556/2778 | 1.62 (1.46 to 1.79) | <0.001 |
| Only negative swabs (Y/N) | 0 | 1588/1746 | 1.03 (0.94 to 1.12) | 0.579 |
| Mean no symptoms /week | 9 | 0.1 (0.0-5.0) | 2.03 (1.91 to 2.15) | <0.001 |
| SARS-CoV-2 vaccine | 0 | 1268/2066 | 0.90 (0.82 to 0.99) | 0.035 |

BMI, Body Mass Index; Y, Yes; N, No; CI, Confidence Interval

**FIGURES (Supp.)**

Figure S1. Study timeline in relation to the number of Severe Acute Respiratory Syndrome Coronavirus-2 (SARS-CoV-2) cases in Switzerland (source: Swiss Federal Office of Public Health).

Figure S2. Complete case analysis for the sum score of the post-concussion (RMEAD), fatigue severity (FSS), depression (PHQ), and anxiety (GAD) scores by healthcare workers (HCW) with positive SARS-CoV-2 swab (red), seropositive HCW without positive swab (orange), and seronegative HCW without positive swab (light blue). Statistics: Positive HCW are compared to negative HCW, respectively (*** p-value <0.001; ** p-value 0.001-0.01; * p-value 0.01-0.05).

Figure S3. Sum score for the post-concussion (RMEAD), fatigue severity (FSS), depression (PHQ), and anxiety (GAD) scores according to weeks since positive swab, compared to negative controls.

Figure S4. Forest plot showing results of multivariable Poisson regression analysis on the multiplicative effect on the score of the Rivermead Post Concussion Symptoms Questionnaire (RMEAD, left) and Fatigue Severity Scale (FSS, right) score. Factors in blue are negatively, those in red positively associated with the presence of symptoms.
